# Characterization of SARS CoV-2 Antibodies in Breast Milk from 21 Women with Confirmed COVID-19 Infection

**DOI:** 10.1101/2021.07.19.21260661

**Authors:** Lars Bode, Kerri Bertrand, Julia A. Najera, Annalee Furst, Gordon Honerkamp-Smith, Adam D. Shandling, Christina D. Chambers, David Camerini, Joseph J. Campo

## Abstract

One potential mechanism for protection from SARS-CoV-2 in infants and young children is through passive immunity via breast milk from a mother previously infected with the novel coronavirus. The primary objectives of this study were to establish the presence of SARS-CoV-2 specific IgA and IgG and to characterize the specific antigenic regions of SARS-CoV-2 proteins that were reactive with antibodies in breast milk from women with confirmed SARS-CoV-2 infection.

Between March 2020 and September 2020, 21 women with confirmed SARS-CoV-2 infection were enrolled into Mommy’s Milk at the University of California, San Diego. Women donated serial breast milk samples. Breast milk samples were used to probe a multi-coronavirus protein microarray containing full-length proteins and variable length overlapping fragments of SARS-CoV-2 spike (S), envelope (E), membrane (M), nucleocapsid (N), and open reading frame (ORF) proteins.

The breast milk samples contained IgA reactive with a variety of SARS-CoV-2 antigens. The most IgA-reactive SARS-CoV-2 proteins were N (42.9% of women responded to 1 ≥ N fragment) and S proteins (23.9% of women responded to ≥ 1 fragment of S1 or S2). Overall, individual COVID-19 cases had diverse and unique milk IgA profiles over the course of follow-up since onset of SARS-CoV-2 symptoms.

## Introduction

To date, the novel severe acute respiratory syndrome coronavirus 2 (SARS-CoV-2) has infected over 33.5million people in the United States, resulting in over 607,000 deaths in less than 19 months (1). Although COVID-19, the disease caused by SARS-CoV-2, is typically mild in children compared with adults, severe disease and death have been reported in newborns, infants and young children (2, 3). Multisystem inflammatory syndrome in children (MIS-C) can occur even after resolution of infection, and has been disproportionately affecting minority ethnic children (4). Therefore, protecting the vulnerable infant and toddler population from SARS-CoV-2 infection is critical. One potential mechanism for protection is through passive immunity via breastfeeding from a mother previously infected with SARS-CoV-2.

It is well documented that breast milk contains antibodies in response to viral infections (5–9). It provides passive immunity via immunoglobulins and other bioactive factors such as lactoferrin. Approximately 90% of the antibodies found in breast milk are IgA and 8% are IgM, predominately in secretory form (sIgA/sIgM), which helps protect the antibodies from the harsh environments of the infant mouth and gut. The remaining 2% of antibodies are IgG, which are derived from serum (10).

Few studies have examined the presence of antibodies to SARS-CoV-2 in breast milk. Early in the pandemic, Dong et al. (2020) confirmed the presence of both IgA and IgG antibodies to the SARS-CoV-2 spike (S) protein in breast milk from one previously infected woman, and Favara et al. (2020) found both IgA and IgG antibodies against SARS-CoV-2 nucleocapsid (N) protein, S protein in general, and the receptor binding domain (RBD) of S in particular, in the milk of another previously infected breastfeeding woman (11, 12). Similarly, Fox et al. (2020) reported that milk from 12 of 15 infected women contained IgA that was reactive to the RBD of the SARS-CoV-2 spike protein (13). The largest study to date examined 37 breast milk samples from 18 infected women (14). Pace et al. (2021) found that 76% of the milk samples contained SARS-CoV-2-specific IgA and 80% had SARS-CoV-2 specific IgG. In addition, the concentrations of SARS-CoV-2 IgA were consistently higher than those of IgG, which confirms the earlier report of Favara et al. (12, 14). Gao et al. (2020) was the first to report on the presence of SARS-CoV-2 IgM in milk samples from 3 of 4 (75%) infected women in their study (15). However, IgA was not measured in these samples. None of these studies examined the differences in antibody profiles between women with varying levels of COVID-19 severity.

The primary objectives of this study were to establish the presence of SARS-CoV-2 specific IgA and IgG and to characterize the specific antigenic regions of SARS-CoV-2 proteins that were reactive with antibodies in breast milk from women with confirmed SARS-CoV-2 infection.

## Results and Discussion

### Maternal Characteristics

Between March 14, 2020 and September 1, 2020, 21 women who had a confirmed SARS-CoV-2 infection by RT-PCR were enrolled in Mommy’s Milk Human Milk Research Biorepository at UC San Diego. As shown in Table 1, the majority of women were White (85.7%) and all non-Hispanic. Maternal age at enrollment averaged 34.49 years (SD 3.67) and child age at enrollment averaged 10.17 months (SD 5.45). Five of the women (23.8%) had a body mass index (BMI) ≥ 30, and 9 women (42.9%) had underlying health conditions, including asthma, hypertension, diabetes, heart defects or heart conditions, kidney defects or kidney conditions, hypothyroidism, hyperthyroidism, or irritable bowel disease. All women were symptomatic for COVID-19, two of whom were hospitalized. The number of symptoms presented on average were 9.57 (SD 4.02) and symptoms lasted an average of 25.14 days (SD 15.82). Milk samples were collected at time of onset of symptoms, and an additional one to twelve samples were taken at a range of 1 to 231 days post-onset of symptoms (mean number of samples collected per woman: 4.76 (SD 2.95; Range 2-13); mean number of days of continuation of symptoms: 25.14 days (SD 15.82, Range 10-91), days.

**Table 1.** Characteristics of Women who Tested Positive for SARS-CoV-2 and their Breastfed Children, N=21 women and 22 children.

| Select Characteristics | n % |
| --- | --- |
| Maternal Age – years (SD) | 34.49 (3.67) |
| Race – n (%) |  |
| Caucasian | 18 (85.7) |
| Asian | 2 (9.5) |
| Other | 1 (4.8) |
| Ethnicity – n (%) |  |
| Non- Hispanic | 21 (100.0) |
| US Region of Residence – n (%) |  |
| North-East | 7 (33.3) |
| South | 5 (23.8) |
| Mid-West | 5 (23.8) |
| West | 4 (19.0) |
| BMI $\geq 30$ – n (%) | 5 (23.8) |
| Underlying Health Condition<sup>1</sup> - n (%) | 9 (42.9) |
| Symptomatic – n (%) | 21 (100.0) |
| Hospitalized – n (%) | 2 (9.5) |
| Treatment with Remdesivir – n (%) | 1 (4.8) |
| Treatment with ECMO – n (%) | 1 (4.8) |
| Number of Symptoms-mean (SD), [range] | 9.57 (4.02), [1,16] |
| Number of Days Symptomatic – mean (SD), [range] | 25.14 (15.82), [10, 91] |
| Infant Age (months)–mean(SD) | 10.17 (5.45) |
| Infant Sex – n (%) |  |
| Male | 11 (50.0) |
| Female | 11 (50.0) |
| Pre-Term Delivery – n (%) | 5 (23.8) |

<sup>1</sup>Underlying health conditions included: Asthma, Hypertension, Diabetes, Heart Defect/ Conditions, Kidney Defect/ Condition, Hypothyroidism, Hyperthyroidism, IBD/ IBS

**Table 2.**
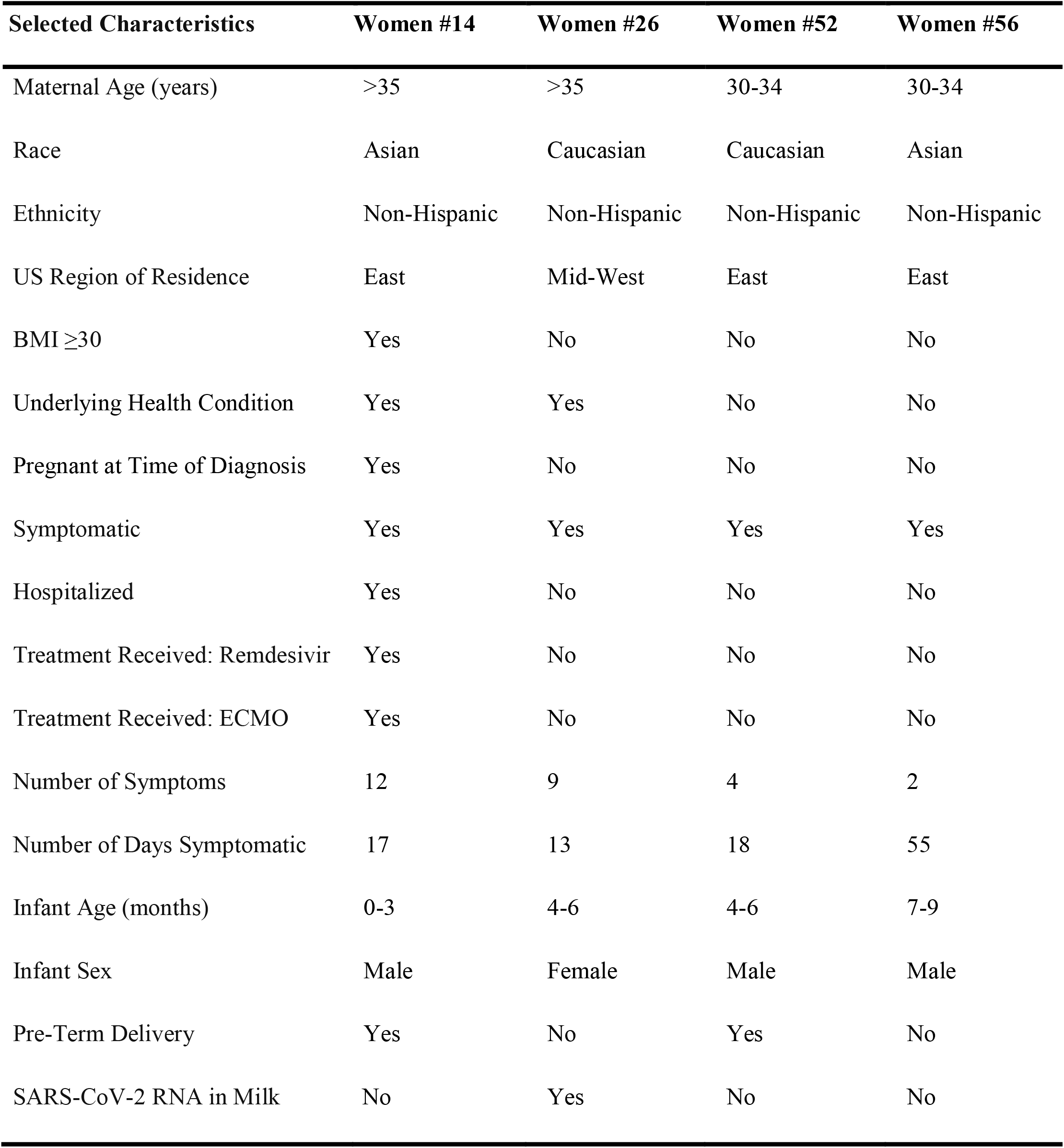
Specialized Profiles for Women #14, 26, 52, 56 with unique antibody response to SARS-CoV-2.

| **Selected Characteristics** | **Women #14** | **Women #26** | **Women #52** | **Women #56** |
| --- | --- | --- | --- | --- |
| Maternal Age (years) | >35 | >35 | 30-34 | 30-34 |
| Race | Asian | Caucasian | Caucasian | Asian |
| Ethnicity | Non-Hispanic | Non-Hispanic | Non-Hispanic | Non-Hispanic |
| US Region of Residence | East | Mid-West | East | East |
| BMI $\geq 30$ | Yes | No | No | No |
| Underlying Health Condition | Yes | Yes | No | No |
| Pregnant at Time of Diagnosis | Yes | No | No | No |
| Symptomatic | Yes | Yes | Yes | Yes |
| Hospitalized | Yes | No | No | No |
| Treatment Received: Remdesivir | Yes | No | No | No |
| Treatment Received: ECMO | Yes | No | No | No |
| Number of Symptoms | 12 | 9 | 4 | 2 |
| Number of Days Symptomatic | 17 | 13 | 18 | 55 |
| Infant Age (months) | 0-3 | 4-6 | 4-6 | 7-9 |
| Infant Sex | Male | Female | Male | Male |
| Pre-Term Delivery | Yes | No | Yes | No |
| SARS-CoV-2 RNA in Milk | No | Yes | No | No |

### Breastfeeding mothers vary in the production of milk IgA against SARS-CoV-2 proteins and recognition of specific antigenic regions

Breast milk samples from the subjects in this study that were probed on a multi-coronavirus protein microarray (Table S1) and using the electro-chemiluminescent immunoassay (ECLIA) contained IgA reactive with a variety of SARS-CoV-2 antigens as well as antigens from other human coronaviruses (Fig. 1 and 2). A total of 24 SARS-CoV-2 full-length or fragmented proteins expressed using an *E. coli* cell-free *in vitro* transcription and translation system (IVTT) had IgA reactivity above the reactivity threshold of 1.0 normalized log2 signal intensity (twice the background levels) in at least one of the study patients. The most IgA-reactive SARS-CoV-2 proteins were N (9/21 responded to at least one N fragment) and S proteins (5/21 responded to at least one fragment of S1 or S2), although one patient exhibited strong IgA reactivity with M protein. Seropositivity rates to purified recombinant proteins on the arrays and proteins on the ECLIA platform were higher: 19/21 for N by ECLIA, 18/21 for S by both protein array and ECLIA, only 2/21 for the RBD by array but 16/21 by ECLIA, and 12/21 for the NTD by ECLIA. One patient responded to the SARS-CoV-2 nonstructural protein ORF3a, and one other patient responded to ORF7a, although moderate levels of IgA binding (approximately 0.5 normalized signal intensity) was observed against ORF7a in many of the women (Fig. 2). IgG seropositivity rates for IVTT proteins and array purified proteins were similar to IgA, but with more N responders (12/21), whereas N responders by ECLIA were fewer than for IgA (12/21) (Fig. S1). These profiles contrast with our recent observations of SARS-CoV-2 specific IgG in symptomatic COVID-19 patients, where most patients responded to the SARS-CoV-2 S or N proteins with the exception of a minor subset of “serosilent” or “serodelayed” patients (16–18). However, the levels of reactivity seen in this study are in agreement with recent studies of maternal serum and cord blood IgG in SARS-CoV-2 infected pregnant women, where 65% of infected women responded to the RBD and 70% to the N protein (19).

**Figure 1.**
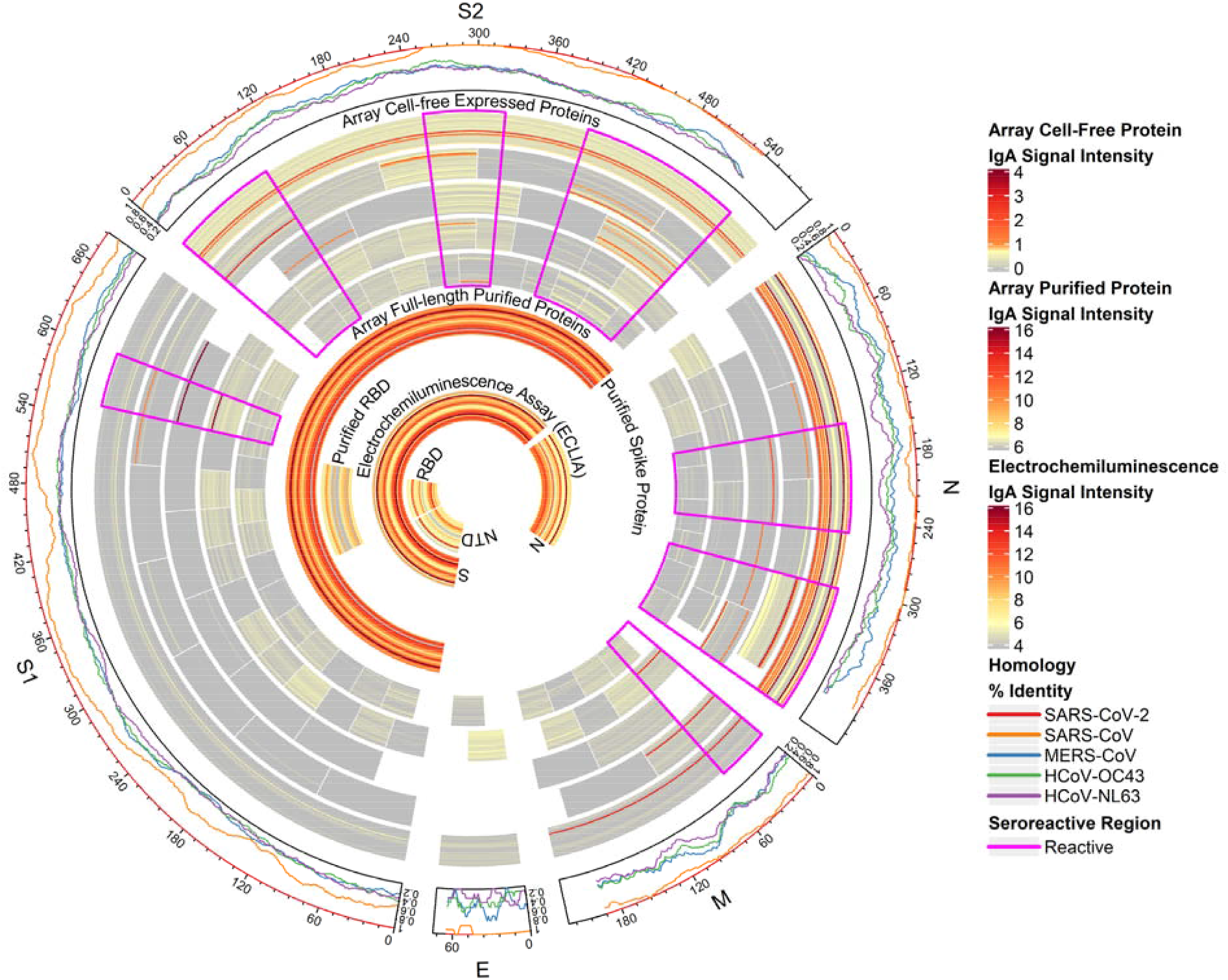
Reactivity of individual COVID-19 patient breast milk IgA to SARS-CoV-2 proteins. The circular graphic maps the amino acid (aa) position of SARS-CoV-2 fragments, showing a heat map of antibody levels for each individual mother for overlapping regions of different aa length. Proteins are indicated outside the circle plot above an axis that shows aa positions from the N-terminus to C-terminus of each protein. The following line graph shows the sequence homology of other HCoVs with SARS-CoV-2 for each gene. The inner circular heat map shows proteins and protein fragments produced in cell-free *E. coli in vitro* transcription and translation reactions by bars that represent length and position of each fragment in each protein. Full-length, 100 aa and 50 aa fragments are shown. Fragments of 30 aa size were mostly non-reactive and are not shown, but are included in the full data sets (See Supplemental Data). Each fragment is drawn 21 times, once for each mother, and shows the maximum normalized signal intensity (SI) of antibody binding to each fragment, for COVID-19 maternal patients among all of their respective breast milk samples taken after onset of symptoms. IgG signal intensity is shown by color gradient (grey to red). Seroreactive regions of the proteins are highlighted by magenta outlines. The inner circle bands represent the responses to full-length purified recombinant S protein (shown crossing both S1 and S2 regions) and the receptor binding domain (RBD) of S protein from the array. This is followed by responses acquired in the electrochemiluminescence assay (ECLIA) to the full-length S and N proteins, the N-terminal domain (NTD) of S protein and the RBD of S protein.

**Figure 2.**
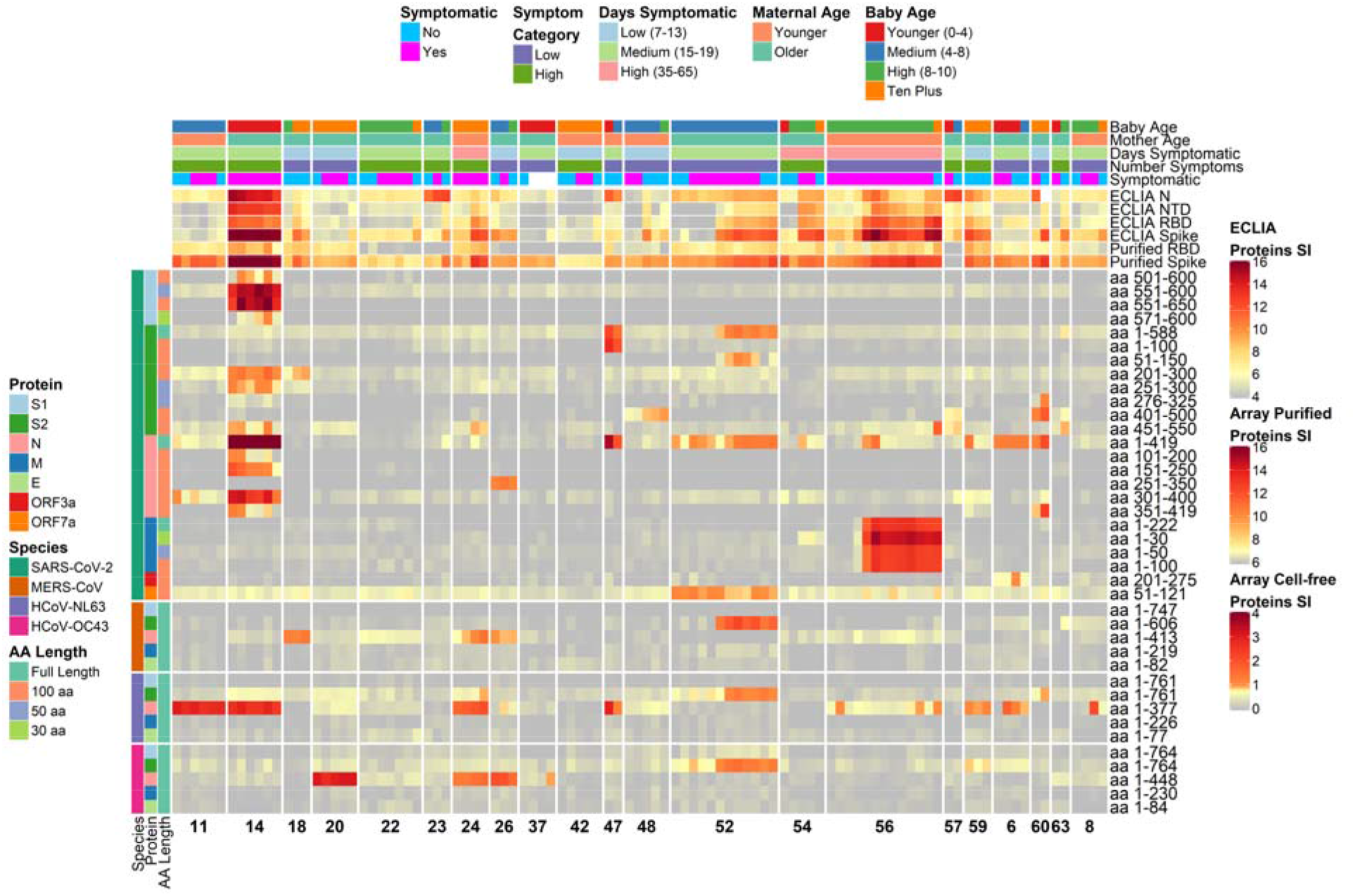
Heatmap depicting relative IgA antibody responses to SARS-CoV-2 as compared to other HCoVs and clinical data. The heatmap presents the signals of antibody binding to individual proteins and protein fragments within the antigenic regions of SARS-CoV-2, as well as the full-length structural proteins of MERS-CoV, HCoV-NL63 and HCoV-OC43, for individual samples. Columns represent breast milk samples, and rows represent proteins or protein fragments: 26 SARS-CoV-2 proteins or fragments filtered for having a maximal normalized log2 signal intensity of at least 0.5 in one or more mother’s samples, and five proteins each of MERS-CoV, HCoV-OC43 and HCoV-NL63. Antibody signal intensity is shown on a color scale from grey to red. Log2 signal intensities from recombinant purified proteins on the array and log2 signal intensity from proteins assayed on the ECLIA platform are overlaid above the array cell-free expressed proteins and shown with independent grey-to-red color scales. Sample clinical information is overlaid above the heatmaps and includes categories at time of sampling for COVID-19 symptoms, number of symptoms, number of days symptomatic, maternal age and baby age. Protein/fragment information is annotated to the left of the heatmaps and includes the virus, full-length protein name and the amino acid length of the protein fragments (“AA Length”, as full length, 100, 50 or 30 aa).

Responses to reactive regions of SARS-CoV-2 structural proteins outlined in the circular map (Fig. 1) were heterogeneous. For example, the C-terminal region of S1 spanning aa 551-650 was recognized by only subject 14 (Fig. 2). Subjects 47 and 52 had milk IgA that recognized the full-length S2 IVTT protein, but subject 47 reacted with a fragment spanning aa 1-100, whereas subject 52 reacted with a fragment spanning aa 51-150 with identical kinetics to full-length S2. Both subjects responded to what are likely unique epitopes within the first 150 aa of S2. Subject 14, however, responded to neither of the first 150 aa fragments of S2 nor the full-length protein, but responded specifically in the regions of aa 201-300 and aa 451-550. Subject 60 had unique IgA reactivity to the aa 401-500 fragment, and subject 48 had borderline reactivity to this fragment as well. For N protein, subject 14 responded to multiple fragments, whereas most N-seropositive subjects responded only to the full-length protein. Subject 26 had a seropositive IgA response only to the aa 251-350 fragment of N protein, which was non-reactive in all other subjects. The IgG levels to SARS-CoV-2 proteins measured in milk from the women in this study were similarly heterogeneous (Fig. S1). In another recent study, variation in the SARS-CoV-2 IgG and IgA responses in COVID-19 mRNA-vaccinated or infected pregnant and lactating women was higher in the milk than in the serum (20). Although antibodies were only tested against the RBD, the study by Collier et al (2021), along with our recent profiling studies in COVID-19 patient sera, indicate that the systemic response to SARS-CoV-2 measured in serum may be less variable than the mucosal response in breast milk (16, 17).

### Diverse IgA kinetic profiles among lactating COVID-19 patients

Individual COVID-19 cases had interesting and unique milk IgA profiles over the course of follow-up since onset of symptoms (Fig. 3). The mother with subject ID #14 had the highest IgA levels among all of the mothers, and IgA levels against both protein array antigens and ECLIA proteins were high from the initial milk sample taken. In particular, mother #14 was the only responder to the C-terminal region of S1 spanning aa 551-650, and she was also the strongest responder to the full-length N protein. This patient was tested positive for SARS-CoV-2 during the end of her pregnancy and had a pre-term delivery. The early high levels of IgA antibodies suggests an anamnestic response, perhaps due to cross-reactivity with other human coronaviruses. Indeed, this patient had a high response to HCoV-NL63 N protein, however, she did not respond to NL63 S2 protein. It is possible that the patient began having symptoms at a later stage of the SARS-CoV-2 infection, thereby missing the early rise in antibodies. Due to the short follow-up period of 7 days after onset of symptoms, it is not clear if IgA levels would contract, as seen with some other patients.

**Figure 3.**
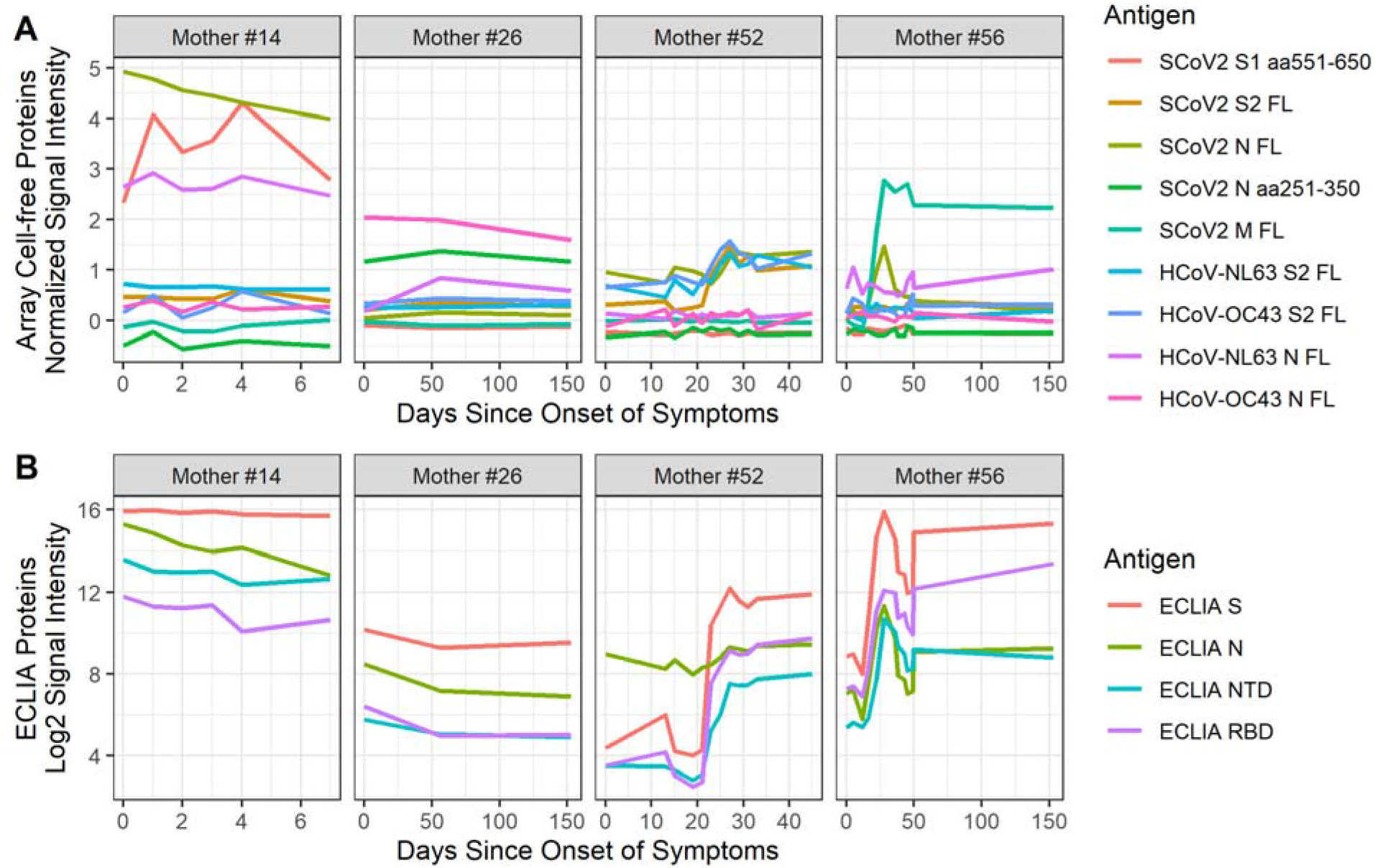
Unique longitudinal profiles of mothers’ breast milk IgA response to SARS-CoV-2 show heterogeneity in antibody recognition. (**A**) The line plots show breast milk IgA responses to selected full-length and protein fragments produced by the cell-free *E. coli* in vitro transcription and translation system. Antigens were selected to illustrate differences in reactivity profiles of four unique responders to SARS-CoV-2. The timing of breast milk sampling in days since symptomatic is shown on a free x-axis, and the normalized IgA signal intensity is shown in the y-axis. Each colored line represents the longitudinal samples of one of the four mothers. S2 and N proteins from the common cold human coronaviruses HCoV-NL63 and HCoV-OC43 are shown in the second row of panels. aa: amino acid; FL: full-length. (**B**) The line plots show the log2 signal intensity from proteins assayed on the ECLIA platform, where each panel is a unique mother’s responses to the four antigens. The number of days since symptoms when samples were taken is shown on a free x-axis.

Mother #52 exhibited a classical primary immune response kinetic profile with IVTT full-length S2 and N proteins, beginning low and rising above the seropositivity threshold at approximately 25 days post-onset of symptoms, which also tracked with NL63 and OC43 S2 proteins but not N protein. For this patient, it is possible that symptoms began early in the course of SARS-CoV-2 infection. Mother #56 also had a classical primary immune response kinetic profile, with IgA levels peaking at around 25 days post-onset of symptoms, followed by a moderate contraction, particularly with IVTT full-length N protein, but also for the ECLIA proteins (Fig. 3) and purified S protein on the array (Fig. 2). This patient had a protracted period of symptoms, lasting 91 days. Most interestingly, mother #56 responded in the same way with very high levels of IgA to SARS-CoV-2 M protein, but not OC43 or NL63 M protein, perhaps because the immunoreactive N-terminal region of M is poorly conserved among these viruses (average sequence identity within the first 50 aa of SARS-CoV-2 M protein using a sliding 30 aa window was 36% for OC43 [range: 30-40%] and 23% for NL63 [23-37%]). Another unique case was the mother with subject ID #26, who was not tested by PCR for SARS-CoV-2 but had a positive PCR result in a breast milk sample. She had a moderate response to SARS-CoV-2 S protein and little else, but responded specifically and solely to a fragment in SARS-CoV-2 N protein spanning aa 251-350. This patient also responded to MERS-CoV N protein (Fig. 2), as well as OC43 N protein, indicating that the seroreactive SARS-CoV-2 N fragment is likely a true positive response. Alignment of the aa sequence of this fragment with the NL63 N protein sequence showed a match of ∼48% sequence identity between the aa 269-328 region of OC43-N protein and aa 257-319 of SARS-CoV-2 N. Alignment of the same region with NL63 N protein showed ∼38% sequence identity between aa 232-307 of NL63 N protein and aa 256-331 of SARS-CoV-2 N, although there was no milk IgA reactivity against NL63 N protein. As interesting as these cases, are the numerous women in this study that responded at low levels or not at all to SARS-CoV-2 antigens with milk IgA or IgG. These patterns together with published studies on systemic immune responses to SARS-CoV-2 illustrate a complex relationship between arms of the immune system during the host response to SARS-CoV-2 infection. The causal link for why one subject responds with milk IgA to different antigenic targets than another subject with the same infection remains unclear, but may be related to preferential presentation of epitopes based on MHC haplotype.

### Association of SARS-CoV-2 protein antibody levels with clinical factors

Our assessment of milk IgA associations with the clinical characteristics of the study patients was limited by the modest sample size of 21 women and the observed heterogeneity in IgA and IgG responses. Presence or absence of symptoms during sampling had no notable effect on antibody levels (Fig. S2 and S3). However, there were outlier antigens with elevated IgA responses for days since onset of symptoms and number of days symptomatic that did not reach statistical significance after correction for false discovery rate. These proteins included primarily fragments of S2 and N proteins, as well as the ECLIA RBD and NTD proteins (Supplemental Data). Maternal age had similar outliers, but which were not significant before P-value adjustment. Other antigens, such as those associated with infant age, had low or negligible levels of IgA reactivity.

This study had several limitations as well as strengths. The collection of breast milk samples were not directly observed and samples were collected with nonstandard sampling time points. Therefore, the breast milk samples collected at the onset of symptoms may not reflect duration of exposure to SARS-CoV-2, which may be attributed for the differences observed in IgA kinetics. We also relied on the maternal report of SARS-CoV-2 test results, symptoms and treatments received, however, all participants completed a semi-structured interview guided by trained study staff who prompted for specifics with the aid of a calendar. Another limitation in our study was lack of reactivity to the S1 protein and its fragments produced *in vitro* using an *E. coli* based reaction mixture. This was likely due to the lack of eukaryotic post-translational modifications, including N-linked glycosylation which is abundant in S1. We compensated for this by including purified S protein and RBD in the array and by assaying purified S, RBD and the NTD of S by electro-chemiluminescent immunoassay (ECLIA). The added value of this study comes from the longitudinal assessment of milk antibody levels and the breadth of antigens covered by the protein microarray and ECLIA platforms, coupled with COVID-19 patient cases with unique profiles.

## Methods

### Participants and Breast Milk Sample Collection

Breast milk samples and clinical information were obtained from women participating in the Mommy’s Milk Human Milk Biorepository at the University of California, San Diego. Women residing in the United States who had a confirmed SARS-CoV-2 infection by RT-PCR were enrolled into the study. Information about demographics, health history, illness and exposure dates, symptoms and SARS-CoV-2 test results were collected by participant interview via telephone. Participants self-collected breast milk samples using a provided collection kit including instructions for expressing and storing their samples. Instructions included hand washing before and after milk expression. Participants who had recovered from their illness at the time of the study interview were asked to ship any frozen samples previously collected at the peak of their symptoms in addition to a fresh milk sample. Fresh samples were shipped on ice within 24 h of collection to the Biorepository and stored at −80°C prior to shipment on dry ice to the Antigen Discovery Institute.

### Protein microarray analysis of breast milk samples

The first generation multi-coronavirus protein microarray, produced by Antigen Discovery, Inc. (ADI, Irvine, CA, USA), included 935 full-length coronavirus proteins, overlapping 100, 50 and 30 aa protein fragments and overlapping 13-20 aa peptides from SARS-CoV-2 (WA-1), SARS-CoV, MERS-CoV, HCoV-NL63 and HCoV-OC43. Proteins and protein fragments were expressed using an *E. coli in vitro* transcription and translation (IVTT) system (Rapid Translation System, Biotechrabbit, Berlin, Germany). Included on the array were four structural proteins and five accessory proteins of SARS-CoV-2: spike (S), which was divided into S1 and S2 regions, envelope (E), membrane (M), nucleoprotein (N), open reading frames (ORFs) 3a, 6, 7a, 8 and 10. Fragments of these nine proteins were made through IVTT in 50% overlapping segments of 30 amino acid (aa), 50 aa and 100 aa lengths. There were also structural proteins produced by IVTT for MERS-CoV, HCoV-NL63 and HCoV-OC43 on the array. Additionally, full-length SARS-CoV-2 S protein and the receptor binding domain (RBD) of S protein were included as purified recombinant proteins, plus overlapping 13-20 aa peptides of the SARS-CoV (2002 SARS epidemic) structural proteins and the S proteins of MERS-CoV, HCoV-NL63 and HCoV-OC43 (Table S1). Purified proteins and peptides were obtained from BEI Resources. SARS-CoV-2 and SARS-CoV S proteins were made in Sf9 insect cells and the SARS-CoV-2 RBD, made in HEK-293 cells. IVTT proteins, purified proteins and peptides were printed onto nitrocellulose-coated glass AVID slides (Grace Bio-Labs, Inc., Bend, OR, USA) using an Omni Grid Accent robotic microarray printer (Digilabs, Inc., Marlborough, MA, USA). Microarrays were probed with breast milk at a 1:15 dilution factor and antibody binding detected by incubation with Cy3 fluorochrome-conjugated goat anti-human IgA (Cat# 109-166-011, Jackson ImmunoResearch, West Grove, PA, USA). A subset of the single most IgA-reactive breast milk sample per patient was also probed at a 1:5 dilution factor for detection of IgG binding using DyLight650 fluorochrome-conjugated goat anti-human IgG (Cat# A80-104D5, Bethyl Laboratories, Inc., Montgomery, TX, USA). Slides were scanned on a GenePix 4300A High-Resolution Microarray Scanner (Molecular Devices, Sunnyvale, CA, USA), and raw spot and local background fluorescence intensities, spot annotations and sample phenotypes were imported and merged in R (R Core Team, 2017), in which all subsequent procedures were performed. Foreground spot intensities were adjusted by subtraction of local background, and negative values were converted to one. All foreground values were transformed using the base two logarithm. The dataset was normalized to remove systematic effects by subtracting the median signal intensity of all IVTT spots for each sample. With the normalized data, a value of 0.0 means that the intensity is no different than the background, and a value of 1.0 indicates a doubling with respect to background. For full-length purified recombinant proteins and peptide libraries, the raw signal intensity data was transformed using the base two logarithm for analysis.

### ECLIA analysis of SARS-CoV-2-specific IgA and IgG in breast milk

V-PLEX COVID-19 Coronavirus Panel 2 multiplex electrochemiluminescence (ECLIA) serology kits were purchased from Meso Scale Discovery (Rockville, MD) to measure Immunoglobulin A and G (IgA and IgG) antibodies against four antigens related to SARS-CoV-2 in whole breast milk samples. Antigens included in panels we analyzed were: SARS-CoV-2 Spike, RBD, N-terminal domain (NTD) of Spike, and N proteins. In brief, pre-coated ECLIA plates were blocked for 1h at room temperature (RT) while shaking with 150 µL Blocker A solution per well (Blocker A solution was prepared according to manufacturer instructions). Plates were washed thrice with 1x MSD Wash Buffer. A 7-point calibration curve with 4-fold serial dilutions and zero calibrator was prepared using stock calibrators using provided Diluent 100. Serum-based reference standard 1, provided with the kits, was used to establish assay calibration curves for each of the antigens. Three control samples, each containing assigned concentrations of each of the antigens were provided by the manufacturer for plate-to-plate assay validation. Whole breast milk samples were diluted 1:10 and 1:100 in 1x PBS for IgG and IgA assays, accordingly. Plates were incubated with 50 µL of prepared calibrators, serology controls, or diluted milk samples for 1h at RT while shaking. Plates were washed thrice with 1x MSD Wash Buffer. Plates were incubated with 150 µL of 1x detection antibody solution containing SULFO-TAG anti-human IgA or IgG prepared with Diluent 100. Plates were washed thrice with 1x MSD Wash Buffer. 150 µL of MSD GOLD Read Buffer B was added to each well, plates read on an ECLIA MESO QuickPlex SQ 120 plate reader, and the generated data analyzed with Discovery Workbench software. ECLIA signals were transformed using the base two logarithm for analysis.

### Statistics

Maternal and infant characteristics were presented as means and standard deviations (SDs). Categorical variables were expressed as counts and percentages. Missing values were excluded. R version 4.1.0 was used for description of maternal and child characteristics.

For protein array and ECLIA results, “reactive antigens” were defined, *post hoc* after observing the heterogeneity in mothers’ responses to SARS-CoV-2 proteins and fragments and modeling negative and positive signal distributions using mixture models (Fig. S4) (21). IgA reactivity for IVTT cell-free expressed proteins was defined as a maximum normalized signal intensity greater than 1.0, equivalent to two times background, at any time point in at least one study participant, which was a more stringent (higher) cutoff than the mixture model cutoffs. IVTT IgG reactivity was defined as normalized signal intensity greater than 2.0, or four times background. Reactivity cutoffs for array purified proteins, peptides and ECLIA proteins for IgA and array peptides IgG were established with the mixture models, whereas arbitrary cutoffs were set for array purified protein and ECLIA IgG signals (Fig. S4D and S4H, respectively).

Clinical variables were associated with SARS-CoV-2-specific IgA antibodies using multivariable linear mixed effects regression (LMER) to model antibody responses to each individual SARS-CoV-2 protein or fragment with random intercepts at the subject level to adjust for repeated measures. LMER models were fit with clinical factors at time of sample collection, including days since symptom onset, presence of COVID-19 symptoms, number of symptoms, number of days symptomatic, maternal age (years) and baby’s age (months) as fixed effects variables. All coefficients were returned from models fit using restricted maximum likelihood (REML). To generate P-values for LMER models, the models were refit using maximum likelihood (ML) and compared by ANOVA against null models with the coefficient removed using ML. For IgG responses, ordinary least squares (OLS) models were fit with clinical factors, since a single sample for each study participant was assayed. P-values were adjusted for the false discovery rate using the method described by Benjamini and Hochberg (22). Data visualization was performed using the circlize, ComplexHeatmap and ggplot2 packages in R (23, 24).

### Study Approval

The University of California San Diego Institutional Review Board approved the study, and women provided oral and written informed consent prior to use of their breast milk samples in the analyses described above.

## Data Availability

Data access may be provided, with appropriate ethics approval, by contacting the authors.

## Author Contributions

Dr. Chambers and Ms. Bertrand participated in study design, supervised the collection of specimens used in the study, and were involved in preparing the manuscript.

Mr. Honerkamp-Smith performed demographic and statistical analysis, and was involved in preparing the manuscript.

Drs. Campo and Camerini and Mr. Shandling participated in study design; developed the methods for and performed breast milk antigen experiments and prepared the manuscript.

Dr. Bode, Dr. Najera and Ms Furst validated the ELCIA assays for use with breast milk and performed breast milk ECLIA assays for IgA and IgG.

Dr. Bode participated in the design of the study and all aspects of data review, provided financial support, and was integrally involved in manuscript finalization.

All authors approved the final manuscript as submitted and agree to be accountable for all aspects of the work.

## Acknowledgements

The authors wish to thank student researchers Angelique Ghadishah, Bianca Kermani, Samantha Powell, and Zoe Sidiropoulos as well as Antigen Discovery Inc. technical staff Arlo Randall and Amit Oberai for helping with data collection and Andy Teng, Chris Hung, Jozelyn Pablo, Vu Huynh and Joshua Edgar for design and fabrication of the multipathogen protein microarray. We also wish to express our appreciation to the women who participated in this study for giving so generously of their time and effort to provide clinical information and breast milk samples.

## Conflict of interest statement

Dr. Bode reported serving as the UC San Diego Chair of Collaborative Human Milk Research, endowed by the Family Larsson-Rosenquist Foundation, which also provided emergency funding for this project. Medela Corporation provided milk sample collection materials for this study. Dr. Chambers reports that shipping of milk samples was financially supported by the Mothers’ Milk Bank at Austin, an accredited milk bank and member of the Human Milk Banking Association of North American, and support for banking of samples in the Mommy’s Milk Human Milk Biorepository at UC San Diego was provided by National Institutes of Health (NIH) National Center for Advancing Translational Sciences (NCATS) under Award Number UL1TR001442 to Gary Firestein at UC San Diego Altman Clinical Translational Research Institute (ACTRI). David Camerini and Joseph J. Campo are employees of Antigen Discovery Inc. The other authors have no relevant conflicts to disclose.

---

**Supplemental Figure 1.**
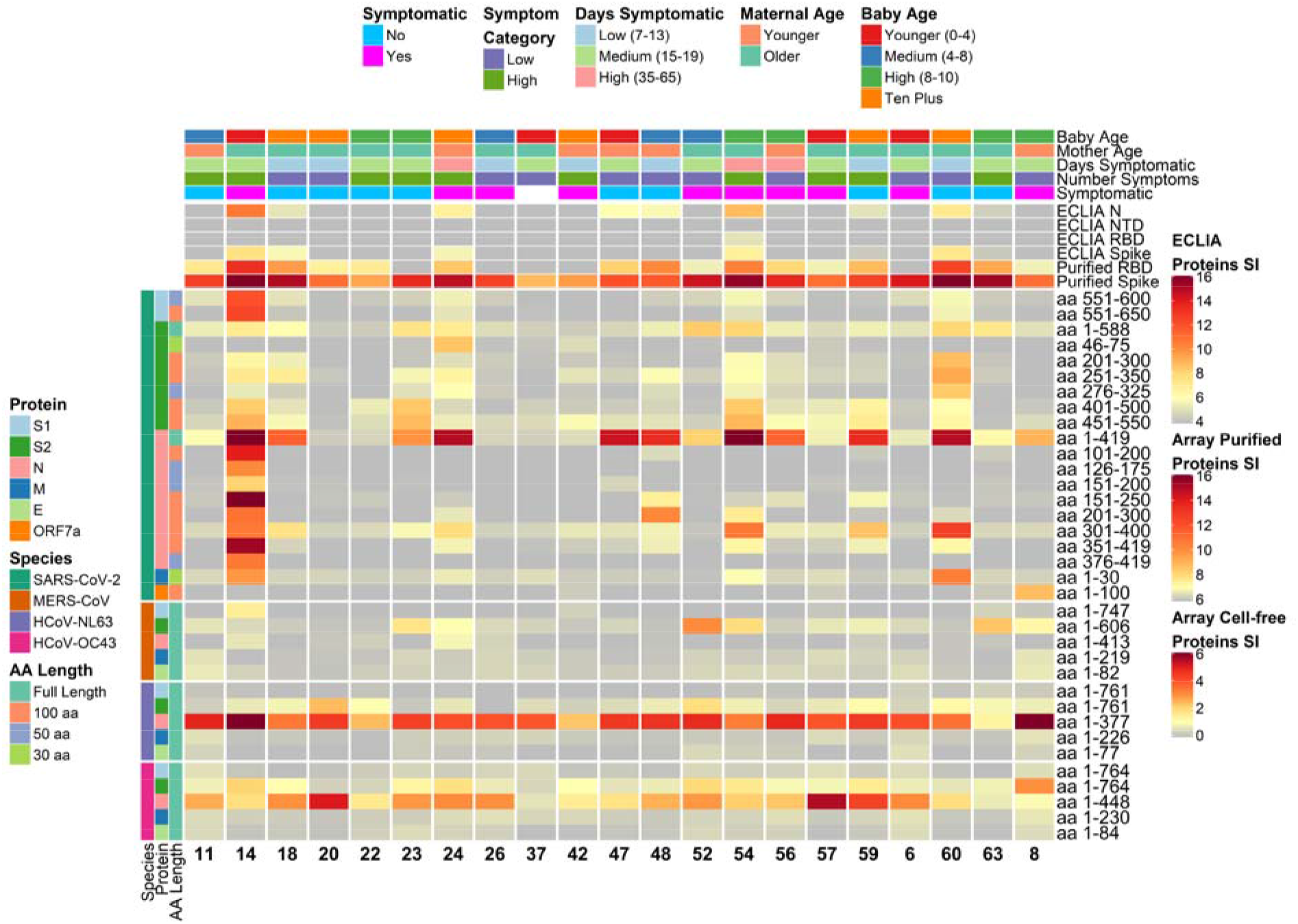
Heatmap depicting relative IgG antibody responses to SARS-CoV-2 as compared to other HCoVs and clinical data. The heatmap presents the signals of antibody binding to individual proteins and protein fragments within the antigenic regions of SARS-CoV-2, as well as the full-length structural proteins of MERS-CoV, HCoV-NL63 and HCoV-OC43, for individual samples. Columns represent breast milk samples that were analyzed for IgG based on selection of one sample per mother with maximal IgA responses. Mother IDs are indicated at the bottom of each column. Rows represent proteins or protein fragments: 20 SARS-CoV-2 proteins or fragments filtered for having a maximal normalized log2 signal intensity of at least 2.0 in one or more mother’s samples (noise levels for breast milk IgG were lower than IgA, and thus the normalized cutoff was set at 2.0 instead of 1.0 used for IgA), and five proteins each of MERS-CoV, HCoV-OC43 and HCoV-NL63. Antibody signal intensity is shown on a color scale from grey to red. Log2 signal intensities from recombinant purified proteins on the array and log2 signal intensity from proteins assayed on the ECLIA platform are overlaid above the array cell-free expressed proteins and shown with independent grey-to-red color scales. Sample clinical information is overlaid above the heatmaps and includes categories at time of sampling for COVID-19 symptoms, number of symptoms, number of days symptomatic, maternal age and baby age. Protein/fragment information is annotated to the left of the heatmaps and includes the virus, full-length protein name and the amino acid length of the protein fragments (“AA Length”, as full length, 100, 50 or 30 aa).

**Supplemental Figure 2.**
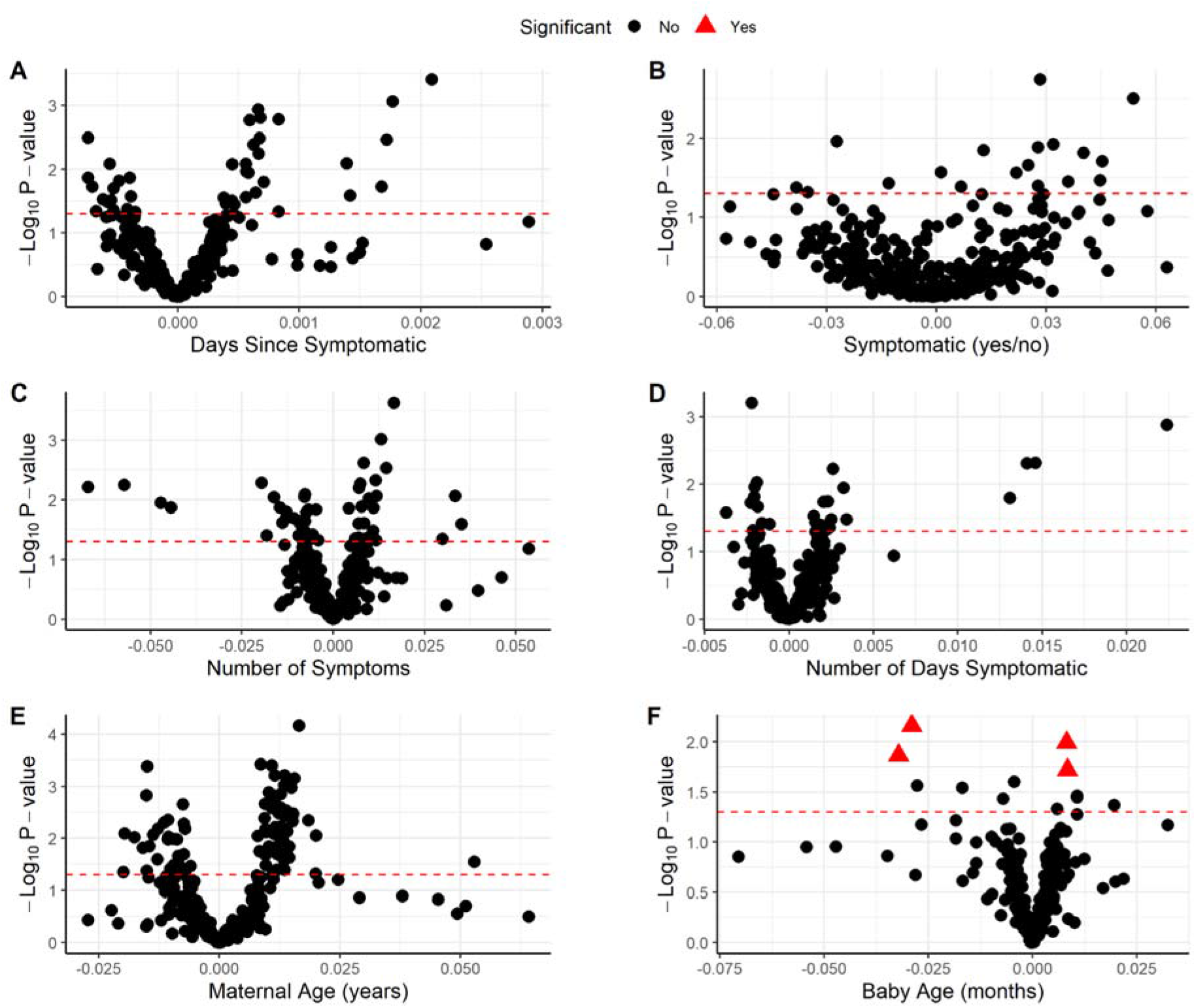
Association of breast milk IgA responses to array cell-free expressed SARS-CoV-2 full length proteins and fragments with clinical factors. The volcano plots show the statistical effect estimates of clinical factors at time of sampling including (**A**) days since onset of symptoms, (**B**) presence of COVID-19 symptoms, (**C**) number of symptoms, (**D**) number of days symptomatic, (**E**) maternal age in years and (**F**) baby’s age in months. The x-axis shows the linear mixed effects regression (LMER) coefficients, and the y-axis shows the inverse log10 p values for each of the SARS-CoV-2 proteins expressed using the cell-free *E. coli* expression system. The proteins/fragments with significant associations after correction for the false discovery rate are highlighted as red triangles.

**Supplemental Figure 3.**
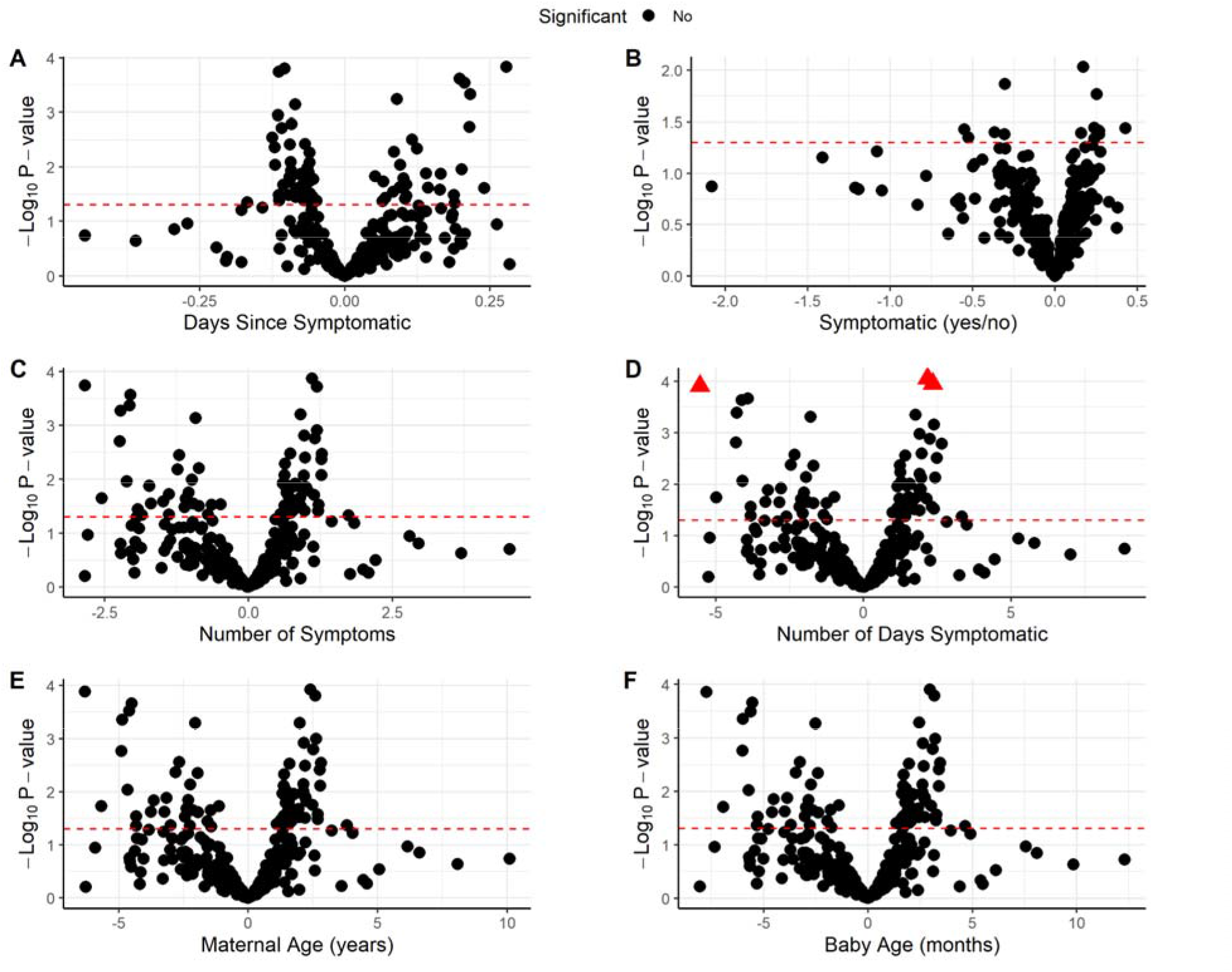
Association of breast milk IgG responses to array cell-free expressed SARS-CoV-2 full length proteins and fragments with clinical factors. The volcano plots show the statistical effect estimates of clinical factors at time of sampling including (**A**) days since onset of symptoms, (**B**) presence of COVID-19 symptoms, (**C**) number of symptoms, (**D**) number of days symptomatic, (**E**) maternal age in years and (**F**) baby’s age in months. The x-axis shows the ordinary least squares (OLS) regression coefficients, and the y-axis shows the inverse log10 p values for each of the SARS-CoV-2 proteins expressed using the cell-free *E. coli* expression system. The proteins/fragments with significant associations after correction for the false discovery rate are highlighted as red triangles.

**Supplemental Figure 4.**
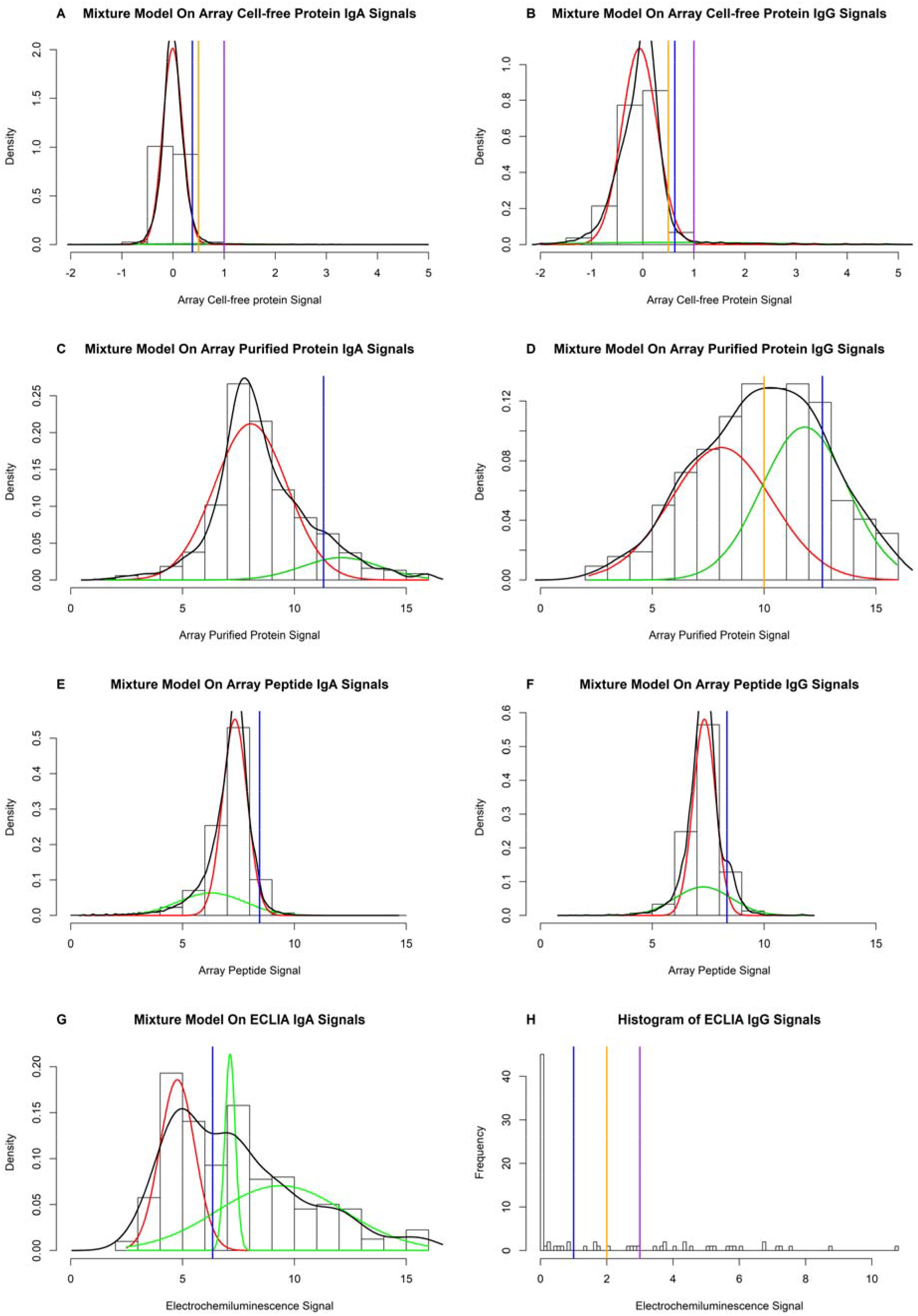
Distribution of milk IgA and IgG signals for array proteins and ECLIA proteins. (**A-G**) Mixture models were fit to antibody signals against different types of proteins on the protein microarray or ELCIA platforms to fit 2-mode Gaussian distributions, except for the ECLIA IgA signals which exhibited a trimodal distribution. Histograms and density curves (black lines) are shown in all plots. The empirically derived distribution of negative signals are shown by red lines, and the “positive” distribution by green lines, except for ECLIA IgA (**G**), which has two green lines for the two positive modes. The blue vertical lines represent seropositivity cutoffs calculated from the mean and 2 standard deviations of the negative distributions. For the array cell-free expressed proteins (**A-B**), cutoffs are also shown for 0.5 (orange vertical line) and 1.0 (purple vertical line). For array purified protein IgG signals (**D**), a bimodal distribution was not observed and the mixture model did not converge, thus a line was drawn at 10.0 (orange vertical line), representing raw median fluorescence intensity (MFI) signals of approximately 1,000 and centered between the negative and positive lines. (**H**) Mixture models did not converge for ECLIA IgG data, thus a histogram is shown with cutoffs drawn at 1 (blue), 2 (orange) and 3 (purple).

